# ERBB4 Drives the Proliferation of BRAF-WT Melanoma Cell Lines

**DOI:** 10.1101/2022.06.20.22276663

**Authors:** L.M. Lucas, R.L. Cullum, J.N. Woggerman, V. Dwivedi, J.A. Markham, C.M. Kelley, E.L. Knerr, L.J. Cook, H.C. Lucas, D.S. Waits, T.M. Ghosh, K.M. Halanych, R.B. Gupta, D.J. Riese

**Author notes:** Corresponding Author: David J. Riese II, Department of Drug Discovery and Development, Harrison College of Pharmacy, Auburn University, Walker 3211g, Auburn, AL. 36849.

## Abstract

Metastatic skin cutaneous melanomas remain a significant clinical problem. In particular, those melanomas that do not contain a gain-of-function *BRAF* allele remain challenging to treat because of the paucity of targets for effective therapeutic intervention. Thus, here we investigate the role of the ERBB4 receptor tyrosine kinase in skin cutaneous melanomas that contain wild-type *BRAF* alleles (“*BRAF* WT melanomas”). We have performed *in silico* analyses of a public repository (The Cancer Genome Atlas - TCGA) of skin cutaneous melanoma gene expression and mutation data (TCGA-SKCM data set). These analyses demonstrate that elevated *ERBB4* transcription strongly correlates with *RAS* gene or *NF1* mutations that stimulate RAS signaling. Thus, these results have led us to hypothesize that elevated ERBB4 signaling which cooperates with elevated RAS signaling to drive *BRAF* WT melanomas. We have tested this hypothesis using commercially available *BRAF* WT melanoma cell lines. Ectopic expression of wild-type *ERBB4* stimulates clonogenic proliferation of the IPC-298, MEL-JUSO, MeWo, and SK-MEL-2 *BRAF* WT melanoma cell lines, whereas ectopic expression of a dominant-negative (K751M) *ERBB4* mutant allele inhibits clonogenic proliferation of these same cell lines. Ectopic expression of a dominant-negative *ERBB4* mutant allele inhibits anchorage-independent proliferation of MEL-JUSO cells and ectopic expression of a dominant-negative *ERBB2* mutant alleles inhibits clonogenic proliferation of MEL-JUSO cells. These data suggest that elevated signaling by ERBB4-ERBB2 heterodimers cooperates with elevated RAS signaling to drive the proliferation of some *BRAF* WT tumors and that combination therapies that target these two signaling pathways may be effective against these *BRAF* WT tumors.

## Introduction

BRAF inhibitors, MEK inhibitors, and immune checkpoint inhibitors (“checkpoint inhibitors”) are transforming the treatment of advanced skin cutaneous melanomas that possess oncogenic *BRAF* mutations (“*BRAF* mutant melanomas”) [1]. A contemporary’ clinical trial reports 34% five-year survival of patients with advanced *BRAF* mutant skin cutaneous melanomas treated with BRAF and MEK inhibitors [2, 3]. Another clinical trial reports 60% five-year survival of patients with advanced *BRAF* mutant skin cutaneous melanomas treated with a combination of immune checkpoint inhibitors [2, 4]. Finally, combining immune checkpoint inhibitors with BRAF and MEK inhibitors will likely lead to further improvements in survival [5].

Unfortunately, approximately 50% of advanced skin cutaneous melanomas possess wild-type *BRAF* alleles, and contemporary treatments of advanced skin cutaneous melanomas that contain wild-type *BRAF* (“*BRAF* WT melanomas”) have yielded less impressive results [1]. In part, these less impressive results are because of a paucity of actionable targets for the effective (targeted) treatment of these tumors [6]. For example, despite the fact that a majority of *BRAF* WT melanomas possess a gain-of-function *RAS* allele or a loss-of-function *NF1* allele, these tumors do not respond to MEK inhibitors [1]. Moreover, the five-year survival of patients with advanced *BRAF* WT skin cutaneous melanomas treated with immune checkpoint inhibitors is only 48%, less than the 60% experienced by patients with advanced *BRAF* mutant skin cutaneous melanomas in a parallel study [4].

Hence, we have attempted to address this gap in treatment efficacy by evaluating the ERBB4 receptor tyrosine kinase as a candidate target in *BRAF* WT skin cutaneous melanomas. Should ERBB4 prove to be a reasonable target in *BRAF* WT skin cutaneous melanomas, we anticipate that strategies that target ERBB4 signaling could be used in combination with immune checkpoint inhibitors to treat these tumors, analogous to what has been proposed for the treatment of *BRAF* mutant skin cutaneous melanomas.

ERBB4 (HER4) is a member of the ERBB family of receptor tyrosine kinases (RTKs), which includes the epidermal growth factor receptor (EGFR), ERBB2 (HER2/Neu), and ERBB3 (HER3). ERBB4 possesses extracellular ligand-binding domains, a single-pass hydrophobic transmembrane domain, an intracellular tyrosine kinase domain, and intracellular tyrosine residues that function as phosphorylation sites. Ligand binding to EGFR, ERBB3, or ERBB4 stabilizes the receptor extracellular domains in an open conformation competent for symmetrical homodimerization and heterodimerization of the receptor extracellular domains. The dimerization of the extracellular domains enables asymmetrical dimerization of the receptor cytoplasmic domains. Phosphorylation of one receptor monomer on tyrosine residues by the tyrosine kinase domain of the other receptor monomer (“cross-phosphorylation”) ensues. This tyrosine phosphorylation creates binding sites for effector proteins and activation of downstream signaling pathways [7].

Elevated signaling by an RTK is a hallmark of many types of cancer. Hence, RTK overexpression, ligand overexpression, and gain-of-function mutations in an RTK gene are all mechanisms for pathologic, elevated RTK signaling. Indeed, EGFR and ERBB2 have been validated as targets for therapeutic intervention in numerous types of tumors; monoclonal antibodies and small molecular tyrosine kinase inhibitors have been approved to treat tumors dependent on these receptors [8–23]. It appears that ERBB3, particularly in the context of ERBB3-ERBB2 heterodimers, also drives various human tumors [24, 25].

In contrast, the role that ERBB4 plays in human tumors remains ambiguous. Part of the ambiguity reflects that an ERBB4 homodimer can function as a tumor suppressor, whereas an ERBB4-EGFR or ERBB4-ERBB2 heterodimer can drive tumor cell proliferation or aggressiveness [7]. Hence, in this work, we attempt to resolve this ambiguity by testing the hypothesis that ERBB4 is sufficient and necessary for the proliferation of *BRAF* WT skin cutaneous melanoma cell lines.

## Results

### A. BRAF WT melanomas do not appear to be less aggressive than BRAF V600X melanomas

The Cancer Genome Atlas – Skin Cutaneous Melanoma (TCGA-SKCM) dataset contains outcome, gene expression, and mutation data for hundreds of skin cutaneous melanomas [26]. We analyzed the TCGA-SKCM dataset to look for meaningful differences between the group of skin cutaneous melanoma patients whose tumors possess *BRAF* WT alleles (“*BRAF* WT melanomas”) and the group of melanoma patients whose tumors have a gain-of-function *BRAF* V600X allele (“*BRAF* V600X melanomas”). **Table 1** compares these two groups.

**Table 1.**
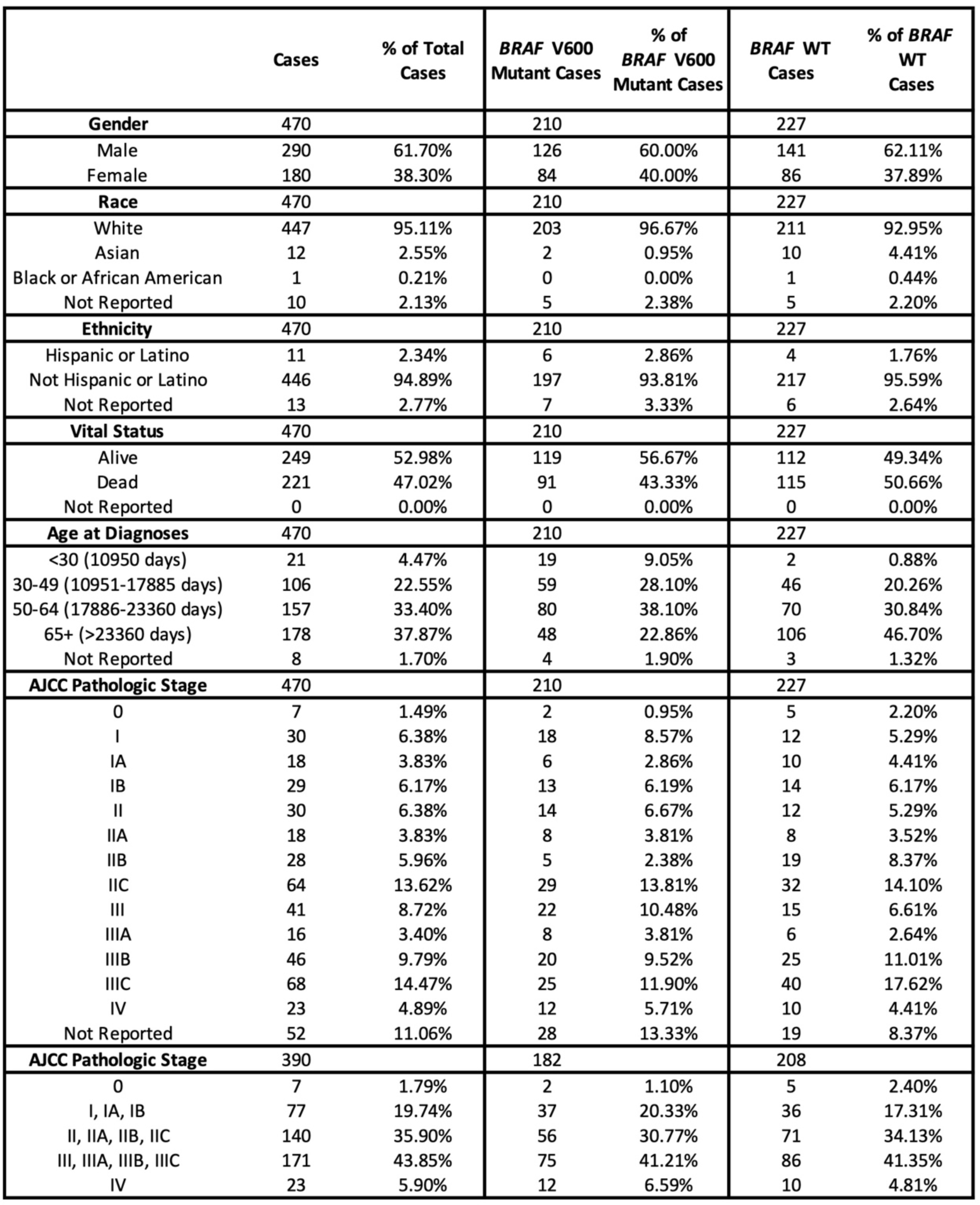
Comparison of demographic and clinicopathologic characteristics of *BRAF* V600X and *BRAF* WT melanoma cases in the TCGA-SKCM dataset.

*BRAF* WT melanomas account for a slightly greater percentage of cases in the TCGA-SKCM dataset than *BRAF* V600X melanomas, suggesting that treating *BRAF* WT melanomas is a significant clinical challenge. Furthermore, Chi-square analysis indicates that a slightly (P=0.1252) greater percentage of *BRAF* WT melanoma patients had died when the dataset was closed than *BRAF* V600X melanoma patients (**Table 2a**). Moreover, Chi-square analysis indicates that the AJCC pathologic stage of the *BRAF* WT melanomas was not significantly different (P=0.6842) from the AJCC pathologic stage of the *BRAF* V600X melanomas (**Table 2b**). Therefore, *BRAF* WT melanomas do not appear to be less aggressive than *BRAF* V600X melanomas. Hence, these *BRAF* WT melanomas pose a significant clinical problem, particularly because there is currently no targeted therapeutic strategy for these tumors.

Gender is associated with disparate melanoma risk and outcomes [27–36]. In particular, it appears that male melanoma patients experience less favorable outcomes. In part, these appear to be due to increased tumor evasion of immunosurveillance and decreased response to immune checkpoint inhibitors. In the *BRAF* WT cases of the ATCC-SKCM dataset, male patients appear to be associated with significantly worse outcomes than female patients (**Table 2c**). However, this gender disparity is not significant among the *BRAF* V600X cases (data not shown).

**Table 2a.** Comparison of survival among *BRAF* V600X and *BRAF* WT melanoma cases in the TCGA-SKCM dataset.

|  | BRAF V600X | BRAF WT | Total |
| --- | --- | --- | --- |
| Alive | 119 | 112 | 231 |
| Dead | 91 | 115 | 206 |
| Total | 210 | 227 | 437 |

$P = 0.1252$

**Table 2b.** . Comparison of AJCC pathological stage among *BRAF* V600X and *BRAF* WT melanoma cases in the TCGA-SKCM dataset.

|  |  | BRAF V600X | BRAF WT | Total |
| --- | --- | --- | --- | --- |
| AJCC Pathologic Stage at Diagnosis | 0 | 2 | 5 | 7 |
|  | I, IA, IB | 37 | 36 | 73 |
|  | II, IIA, IIB, IIC | 56 | 71 | 127 |
|  | III, IIIA, IIIB, IIIC | 75 | 86 | 161 |
|  | IV | 12 | 10 | 22 |
|  | Total | 182 | 208 | 390 |

$P = 0.6842$

**Table 2c.** Association of gender with mortality among the *BRAF* WT melanoma cases in the TCGA-SKCM dataset.

|  | Alive | Deceased | Total |
| --- | --- | --- | --- |
| Female | 50 | 36 | 86 |
| Male | 62 | 79 | 141 |
| Total | 112 | 115 | 227 |

$p = 0.0383$

### B. Elevated ERBB4 expression is correlated with RAS or NF1 mutations in BRAF WT melanomas

Gain-of-function *RAS* gene mutations occur in about 30% of skin cutaneous melanomas, and loss-of-function mutations in *NF1* occur in about 20% of skin cutaneous melanomas. Moreover, gain-of-function *BRAF* mutations, gain-of-function *RAS* gene mutations, and loss-of-function *NF1* mutations are largely mutually exclusive in skin cutaneous melanomas [37].

Receptor tyrosine kinases typically stimulate RAS pathway signaling [38–44]. Hence, we predicted that elevated ERBB4 expression (which is likely to cause elevated ERBB4 signaling) would be inversely correlated with gain-of-function *RAS* gene mutations or loss-of-function *NF1* mutations in *BRAF* WT melanomas of the TCGA-SKCM dataset. *ERBB4* transcription and *NF1*/*RAS* gene expression and mutation data were available for 178 *BRAF* WT melanomas (**Figure 1**). Surprisingly, Chi-square analysis indicates that *ERBB4* transcription greater than or equal to 0.12 (22 melanomas – 12% of the total) is positively correlated (P=0.0057) with a gain-of-function *RAS* gene mutation or a loss-of-function *NF1* mutation in these *BRAF* WT melanomas (**Table 2d**). This correlation suggests that elevated ERBB4 signaling does not stimulate RAS pathway signaling; instead, this correlation suggests that ERBB4 signaling stimulates a pathway that cooperates with elevated RAS pathway signaling to drive *BRAF* WT melanomas.

**Figure 1.**
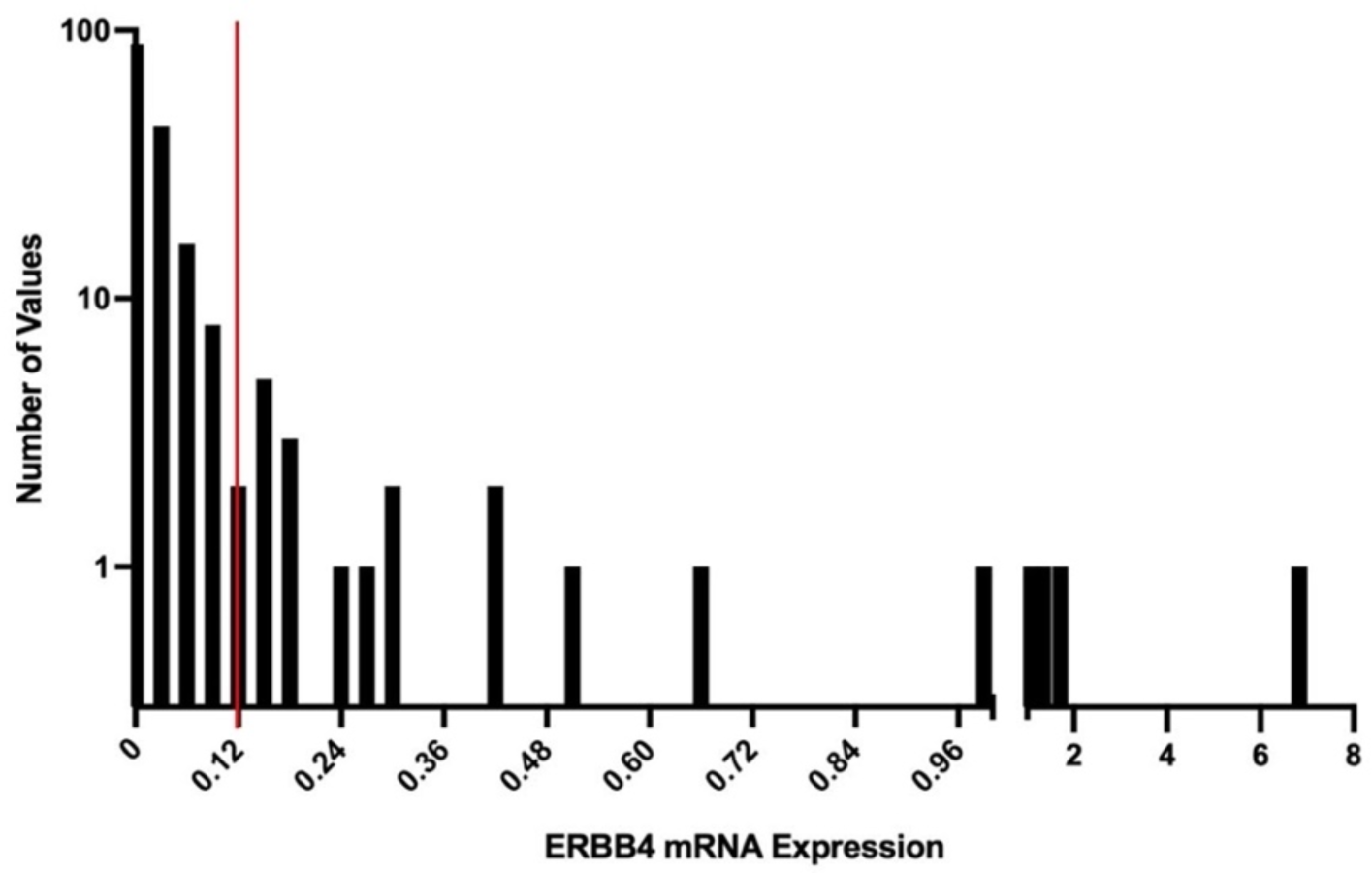
Twenty-two (12%) of the available *BRAF* WT melanomas of the TCGA-SKCM dataset exhibit *ERBB4* transcription of greater than or equal to 0.12.

**Table 2d.** Elevated *ERBB4* transcription is correlated with a gain-of-function *RAS* gene mutation or a loss-of-function *NF1* mutation in *BRAF* WT melanomas of the TCGA-SKCM dataset.

|  | Elevated ERBB4 Expression | Not Elevated ERBB4 Expression | Total |
| --- | --- | --- | --- |
| *RAS* or *NF1* Nonsynonymous Mutation | 21 | 104 | 125 |
| *RAS* and *NF1* WT | 1 | 52 | 53 |
| Total | 22 | 156 | 178 |

$P = 0.0057$

### C. Four commercially available BRAF WT melanoma cell lines appear to be appropriate for analyses of ERBB4 function

A prior report of ERBB4 function in human skin cutaneous melanomas primarily utilized proprietary human skin cutaneous melanoma cell lines [45]. This may have contributed to the failure of others to extend the findings of this work. Hence, we have used the Broad Institute Cancer Cell Line Encyclopedia (CCLE) [46] to identify six commercially available *BRAF* WT melanoma cell lines: COLO-792 [47], HMCB [48], IPC-298 [49], MEL-JUSO [50], MeWo [51], and SK-MEL-2 [52]. The IPC-298, MEL-JUSO, MeWo, and SK-MEL2 cell lines are easily propagated using standard cell culture media and conditions, and our experiments utilize these four cell lines. The IPC-298 and MEL-JUSO cell lines are each derived from the primary melanoma of a female patient, whereas the MeWo and SK-MEL-2 cell lines are each derived from a melanoma metastasis of a male patient (**Table 3**). RNAseq data from the Broad Institute CCLE indicate that these cell lines do not contain gain-of-function mutations in *BRAF* or *PIK3CA*, nor loss-of-function mutations in *PTEN*; however, they do contain driver mutations in *NRAS*, *HRAS*, or *NF1*. Hence, if the malignant phenotypes of these cell lines are dependent on elevated ERBB4 signaling, this elevated ERBB4 signaling may stimulate phosphatidylinositol-3 kinase (PI3K) pathway signaling, which would cooperate with elevated RAS pathway signaling to drive the malignant phenotypes. This hypothesis is supported by our observation that the coupling of ligand-induced ERBB4-EGFR heterodimers to interleukin 3-independent proliferation in BaF3 cells is dependent on ERBB4 coupling to the PI3K pathway [53].

**Table 3.** The commercially available *BRAF* WT melanoma cell lines used in this study do not possess *BRAF*, *PIK3CA*, or *PTEN* mutations, but do possess *NRAS*, *HRAS*, and/or *NF1* mutations. These cell lines exhibit distinct patterns of ERBB receptor gene transcription and ERBB4 ligand gene transcription.

| Cell Line | Gender | Origin | Driver Mutations |  | ERBB4<br>Mutation(s) |
| --- | --- | --- | --- | --- | --- |
|  |  |  | *BRAF/NF1/RAS* | *PIK3CA/PTEN* |  |
| IPC-298 | Female | Primary | *NRAS* Q61L | None |  |
| MEL-JUSO | Female | Primary | *NRAS* Q61L/*HRAS* G13D/*NF1* L1779P | None |  |
| MeWo | Male | Metastasis | *NF1* Q1336*/*NF1* R2053R | None | M766I/S449F |
| SK-MEL-2 | Male | Metastasis | *NRAS* Q61R | None | R50C |

| Cell Line | Transcription of ERBB Receptor Genes |  |  |  | Transcription of Selected ERBB4 Ligand Genes |  |  |  |  |
| --- | --- | --- | --- | --- | --- | --- | --- | --- | --- |
|  | *EGFR* | *ERBB2* | *ERBB3* | *ERBB4* | *NRG1* | *NRG2* | *HBEGF* | *BTC* | *EREG* |
| IPC-298 | 0.07 | 3.58 | 6.18 | <0.01 | 0.03 | 1.36 | 1.76 | <0.01 | 0.01 |
| MEL-JUSO | 0.97 | 3.87 | 5.95 | 0.12 | 3.67 | 2.02 | 6.42 | 0.26 | 0.04 |
| MeWo | 1.43 | 4.74 | 6.59 | 0.69 | 1.01 | 0.28 | 2.05 | 0.46 | 0.03 |
| SK-MEL-2 | 0.06 | 3.18 | 6.74 | 0.01 | 0.77 | 0.73 | 4.49 | <0.01 | <0.01 |

RNAseq data from the Broad Institute CCLE also indicate that these cell lines exhibit different patterns of *ERBB* and ERBB4 ligand gene transcription (**Table 3**). These patterns do not appear to be specific to gender or to primary or metastatic tumors. Furthermore, there does not appear to be any correlation between these patterns of gene expression and the absence or presence of an *ERBB4* mutation.

### D. The canonical ERBB4 JMa/CYT1 isoform appears to be appropriate for studying ERBB4 function in BRAF WT melanoma cell lines

The data presented thus far suggest that ERBB4 signaling drives the proliferation of *BRAF* WT tumors. Thus, we proposed to test whether expression of the wild-type (WT) *ERBB4* gene (cDNA) stimulates the proliferation of *BRAF* WT cell lines and whether expression of a dominant-negative (DN) *ERBB4* mutant inhibits the proliferation of *BRAF* WT cell lines. However, the ERBB4 gene encodes four alternatively-spliced transcripts [7]. The JMa isoforms encode a TACE cleavage site in the extracellular juxtamembrane region, whereas the JMb isoforms lack the TACE cleavage site and flanking amino acids. The CYT1 (CTa) isoforms encode a PPAY amino acid sequence that binds WWOX/YAP proteins; phosphorylation of the tyrosine residue within this sequence creates a binding site for the p85 regulatory subunit of PI3K. The CYT2 (CTb) isoforms lack the PPAY amino acid sequence and flanking amino acids. To establish which *ERBB4* splicing isoform should be utilized in our experiments, we generated the *ERBB4* cDNA from MeWo RNA. We chose MeWo cells for this experiment because they exhibit more *ERBB4* transcription than the other three *BRAF* WT melanoma cell lines (**Table 3**). We used PCR to amplify the region of the *ERBB4* cDNA that distinguishes between JMa and JMb isoforms and the region of the *ERBB4* cDNA that distinguishes between CYT1 and CYT2 isoforms. The size of the JMa/b and CYT1/2 amplicons generated using the *ERBB4* cDNA from the MeWo cells matches the size of the corresponding amplicons generated using an *ERBB4* JMa/CYT1 cDNA plasmid (**Figure 2**). Thus, MeWo cells express the canonical JMa/CYT1 *ERBB4* splicing isoform and it is appropriate to use the canonical JMa/CYT1 *ERBB4* splicing isoform to study *ERBB4* function in MeWo and other *BRAF* WT melanoma cell lines.

**Figure 2.**
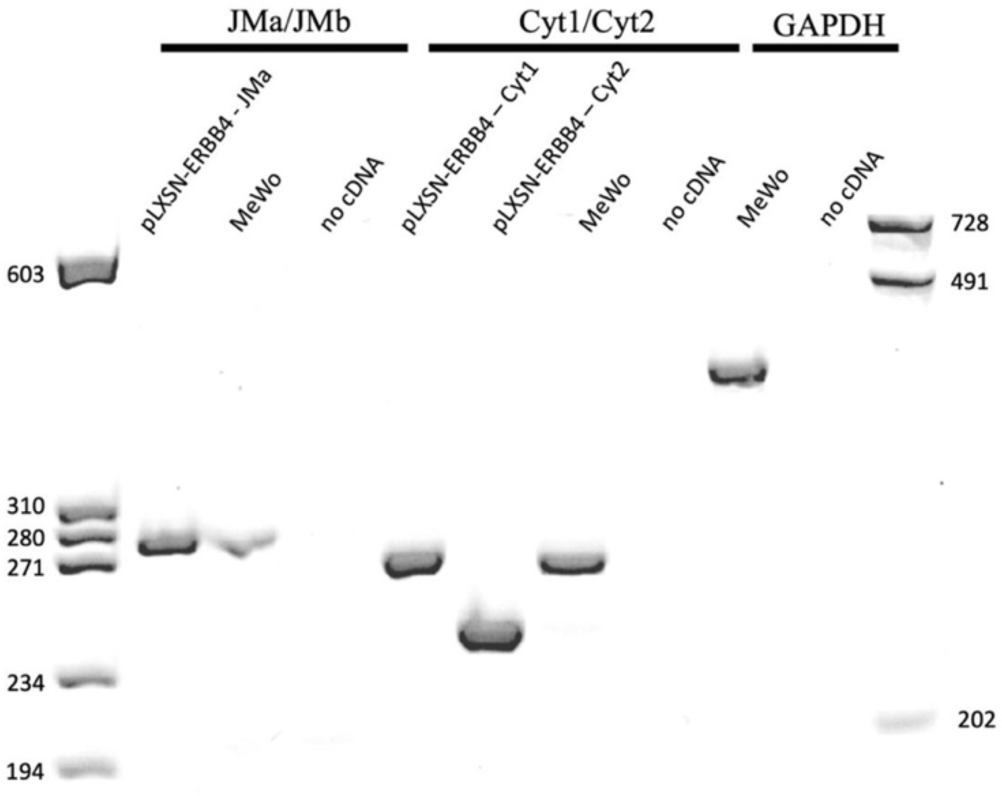
The MeWo cell line expresses the canonical JMa/Cyt1 *ERBB4* transcriptional splicing isoform.

### E. The ERBB4 K751M mutant exhibits decreased tyrosine phosphorylation, enabling it to function as a dominant negative mutant

The EGFR K721A mutant protein lacks tyrosine kinase activity [54], thereby enabling it to function as a dominant negative [55–58] by competitively inhibiting tyrosine phosphorylation by an endogenous, wild-type EGFR molecule. Likewise, the *ERBB2* K753A mutant protein lacks tyrosine kinase activity [59–62], thereby enabling it to function as a dominant negative [62] by competitively inhibiting tyrosine phosphorylation by an endogenous, wild-type ERBB2 molecule. Therefore, we sought to confirm that the analogous ERBB4 K751M mutant protein lacks tyrosine kinase activity and would therefore antagonize signaling by endogenous ERBB4 proteins in BRAF WT melanoma cell lines. We compared ERBB4 protein expression and tyrosine phosphorylation in PA317 cells infected with the LXSN vector control retrovirus, the LXSN-ERBB4-WT retrovirus, and the LXSN-ERBB4-K751M retrovirus (**Figure 3**). PA317 cells that express the ERBB4 K751M mutant protein exhibit markedly less ERBB4 tyrosine phosphorylation than cells that express the wild-type ERBB4 protein. This suggests that the ERBB4 K751M mutant protein lacks tyrosine kinase activity and that the *ERBB4* K751M mutant allele functions as a dominant negative.

**Figure 3.** The ERBB4 K751M mutant protein exhibits much less tyrosine phosphorylation than the wild-type ERBB4 protein.

### F. ERB is sufficient and necessary for clonogenic proliferation of IPC-298, MEL-JUSO, MeWo, and SK-MEL-2 human BRAF WT melanoma cell lines

We have previously used clonogenic proliferation assays to measure the effects of ERBB4 signaling on human prostate [63, 64], breast [64, 65], and pancreatic [66] tumor cell lines. Therefore, we infected IPC-298, MEL-JUSO, MeWo, and SK-MEL-2 *BRAF* WT melanoma cells (**Table 3a**) with a recombinant amphotropic retrovirus that expresses wild-type *ERBB4* (LXSN-ERBB4-WT), a recombinant amphotropic retrovirus that expresses the *ERBB4* K751M dominant-negative mutant allele (LXSN-ERBB4-DN), the vector control amphotropic retrovirus (LXSN), or a mock virus preparation. Because the LXSN recombinant retroviral vector contains a neomycin resistance gene, we selected infected cells using G418. Infection of MEL-JUSO cells with LXSN-ERBB4-WT results in greater clonogenic proliferation than infection with the LXSN control retrovirus. Likewise, infection of MEL-JUSO cells with LXSN-ERBB4-DN results in less clonogenic proliferation than infection with the LXSN control retrovirus (**Figure 4**). Qualitatively similar results were observed when infecting MeWo, IPC-298, and SK-MEL-2 cells (data not shown).

**Figure 4.**
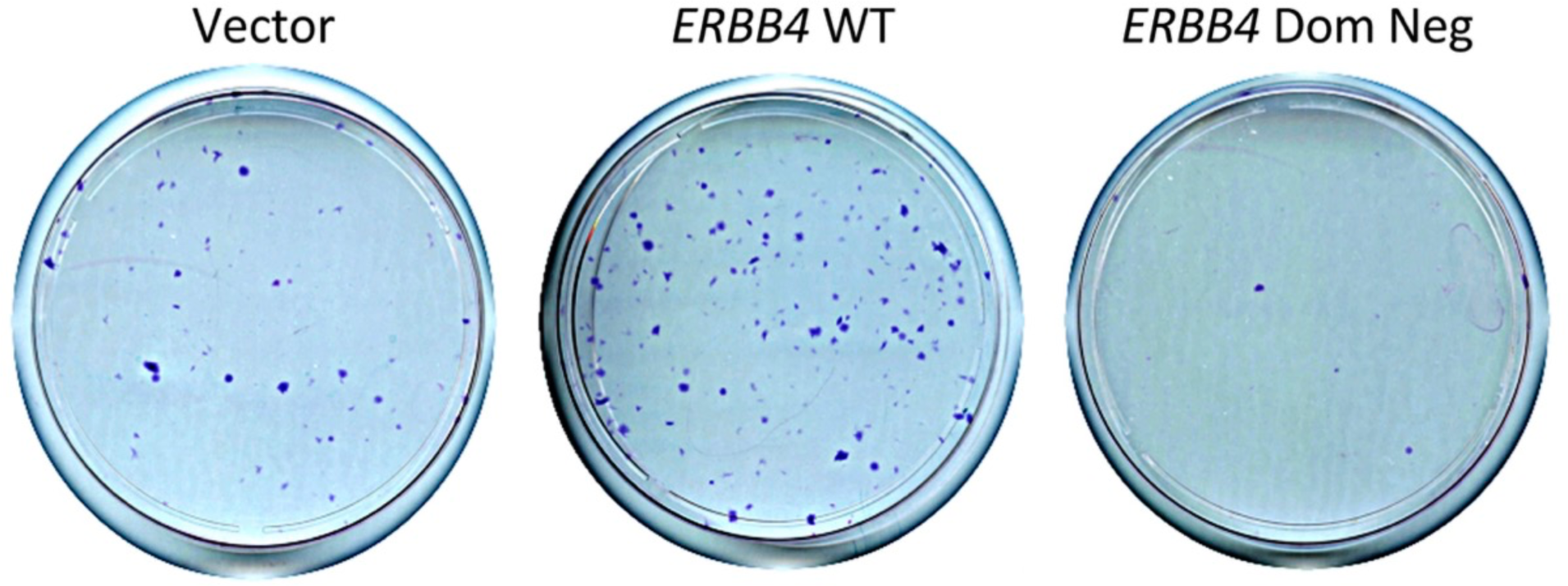
In the MEL-JUSO cell line, stable infection with LXSN-ERBB4-WT causes increased clonogenic proliferation, whereas stable infection with LXSN-ERBB4-DN causes decreased clonogenic proliferation.

To quantitatively assess the effects of *ERBB4* WT and *ERBB4* DN on the clonogenic proliferation of *BRAF* WT melanoma cell lines, in parallel we infected C127 mouse fibroblast cells, which do not endogenously express ERBB4 and do not respond to ERBB4 signaling (data not shown and [63–68]). Relative to the LXSN vector control (100%), *ERBB4* WT causes a modest increase in clonogenic proliferation of SK-MEL-2 (159%) cells, a greater increase in the clonogenic proliferation of MEL-JUSO (389%) and MeWo (563%) cells, and a profound increase in the clonogenic proliferation of IPC-298 (1340%) cells (**Table 4**). All of these increases are statistically significant at p<0.05 (one-tailed).

**Table 4.** In the *BRAF* WT IPC-298, MEL-JUSO, MeWo, and SK-MEL-2 cell lines, *ERBB4* WT causes a statistically significant increase in clonogenic proliferation, whereas *ERBB4* DN causes a statistically significant decrease in clonogenic proliferation.

|  | Efficiency of Clonogenic Proliferation<br>Relative to Vector |  |  |
| --- | --- | --- | --- |
| Cell Line | Vector<br>Control | *ERBB4* WT | *ERBB4* DN |
| IPC-298 | 100% | 1340%<br>n = 5<br>*p* = 0.0256 | 64%<br>n = 5<br>*p* = 0.0082 |
| MEL-JUSO | 100% | 389%<br>n = 9<br>*p* = 0.002 | 35%<br>n = 10<br>*p* = 5.1E-7 |
| MeWo | 100% | 563%<br>n = 5<br>*p* = 0.0173 | 29%<br>n = 5<br>*p* = 0.0003 |
| SK-MEL-2 | 100% | 159%<br>n = 4<br>*p* = 0.0158 | 47%<br>n = 5<br>*p* = 0.0127 |

Likewise, relative to the LXSN vector control (100%), *ERBB4* DN causes a modest decrease in clonogenic proliferation of IPC-298 cells (64%). *ERBB4* DN causes a more pronounced decrease in clonogenic proliferation of the SK-MEL-2 (47%), MEL-JUSO (35%), and MeWo (29%) cells. All of these decreases are statistically significant at p<.05 (one-tailed).

These results suggest that *ERBB4* is both sufficient and necessary for clonogenic proliferation of the four *BRAF* WT melanoma cell lines and that ERBB4 signaling is a driver of *BRAF* WT melanomas.

### F. ERBB4 is necessary for anchorage-independent proliferation of the MEL-JUSO BRAF WT melanoma cell line

We have examined the effects of the *ERBB4* WT and *ERBB4* DN alleles on other characteristics of the MEL-JUSO cell line. Neither the *ERBB4* WT allele nor the *ERBB4* DN allele affects the growth rate or saturation density of the MEL-JUSO cell line (data not shown). Moreover, the *ERBB4* WT allele does not affect the anchorage-independent proliferation of MEL-JUSO cells (**Table 5**). However, the *ERBB4* DN allele causes a decrease in the size of anchorage-independent colonies of MEL-JUSO cells (**Figure 5**). This decrease is statistically significant (Table 5), suggesting that *ERBB4* is necessary for anchorage-independent proliferation of the MEL-JUSO *BRAF* WT melanoma cell line.

**Figure 5.**
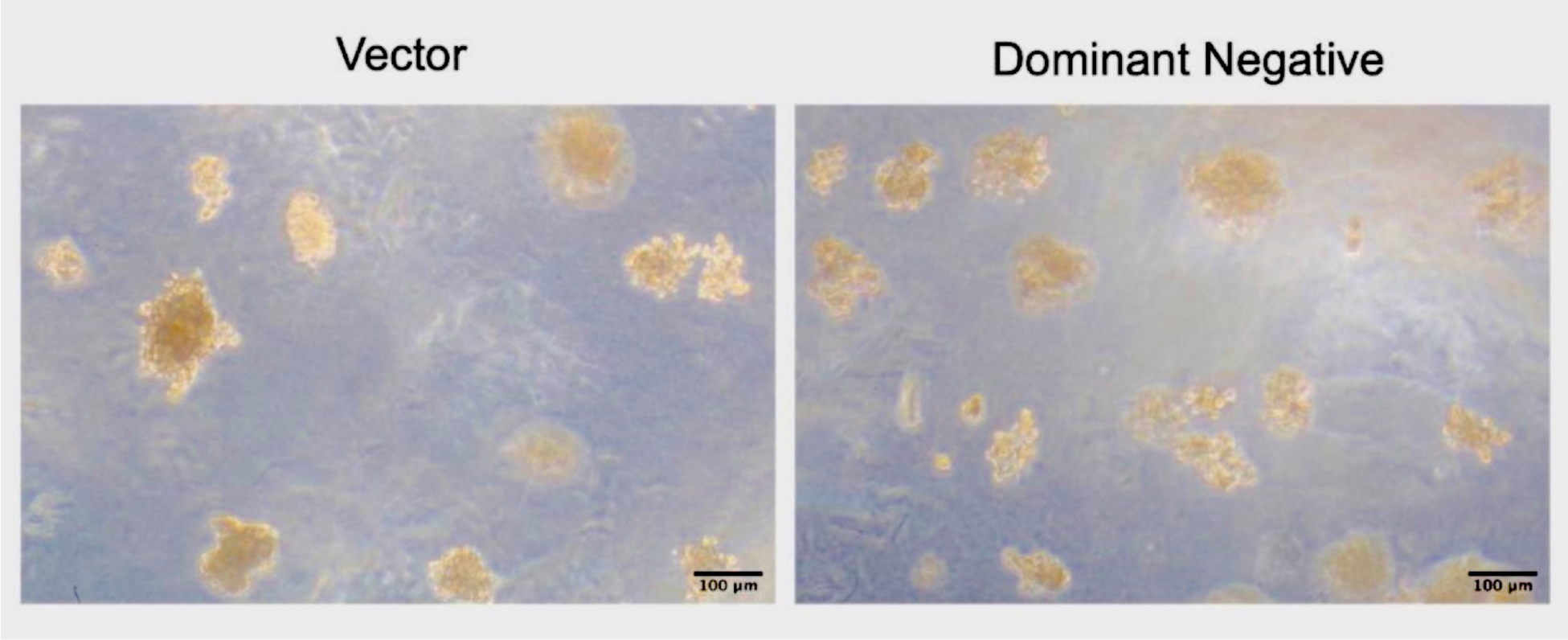
MEL-JUSO cells infected with the LXSN-ERBB4-DN retrovirus form smaller anchorage-independent colonies (right panel) than do MEL-JUSO cells infected with the LXSN vector control retrovirus (left panel).

**Table 5.** *ERBB4* DN causes a significant decrease in the size of anchorage-independent colonies of MEL-JUSO cells.

|  | Vector | WT | DN |
| --- | --- | --- | --- |
| Average (um) | 83 | 83 | 69 |
| SEM (um) | 3 | 3 | 2 |
| Median (um) | 78 | 77 | 63 |
| P-value: T-test<br>(Relative to Vector) |  | 0.483 | 1.932E-04 |
| N (colonies) | 324 | 323 | 326 |

### G. ERBB2, but not EGFR, is necessary for clonogenic proliferation of the MEL-JUSO BRAF WT melanoma cell line

In general, ERBB4 homodimers inhibit cell proliferation, whereas ERBB4-EGFR and ERBB4-ERBB2 heterodimers stimulate cell proliferation [7, 53, 59, 63–66, 68-73]. Therefore, we hypothesized that *EGFR* or *ERBB2* might also be sufficient and necessary for clonogenic proliferation of the MEL-JUSO *BRAF* WT melanoma cell line.

We infected MEL-JUSO and C127 cells with recombinant amphotropic retroviruses that express wild-type *EGFR* (LXSN-EGFR-WT) [59, 71], the *EGFR* K721A dominant-negative (DN) mutant allele (LXSN-EGFR-DN) [59], wild-type ERBB2 (LXSN-ERBB2-WT) [59, 71], the ERBB2 K753A dominant-negative mutant allele (LXSN-ERBB2-DN) [59], wild-type ERBB4 (LXSN-ERBB4-WT) or the LXSN vector control. We quantified clonogenic proliferation (**Table 6**) as described elsewhere. The *EGFR* WT and *ERBB2* WT alleles stimulate clonogenic proliferation of MEL-JUSO cells. Clonogenic proliferation is inhibited by the *ERBB2* DN allele, but not the *EGFR* DN allele. There is no ligand for ERBB2 and the MEL-JUSO cells exhibit transcription of the *NRG1*, *NRG2*, and *HBEGF* genes, all of which encode a ligand for ERBB4 (**Table 3**). Thus, it appears that MEL-JUSO cells exhibit endogenous ligand-induced signaling by ERBB4-ERBB2 heterodimers, which cooperates with elevated RAS signaling to drive the proliferation of these cells.

**Table 6.** In the *BRAF* WT MEL-JUSO cell line, *EGFR* WT and *ERBB2* WT cause an increase in clonogenic proliferation, whereas *ERBB2* DN causes a decrease in clonogenic proliferation.

| Efficiency of Clonogenic Proliferation<br>Relative to Vector |  |  |
| --- | --- | --- |
| *Virus* | *Average*<br>*(n=4)* | *One-Sample*<br>*t Test* |
| LXSN | 100% |  |
| *ERBB4* WT | 233% | 0.064 |
| *EGFR* WT | 244% | 0.011 |
| *EGFR* DN | 144% | 0.485 |
| *ERBB2* WT | 222% | 0.047 |
| *ERBB2* DN | 54% | 0.140 |

## Materials and Methods

### A. Analysis of the TCGA-SKCM dataset

We obtained the following data for all 470 cases of the TCGA-SKCM dataset: gender; race; ethnicity; vital status; age at diagnosis; AJCC pathologic stage at diagnosis; primary tumor site; days to death; copy number variation for ERBB4; mutation status of *BRAF, HRAS, NRAS, KRAS, NF1, EGFR, ERBB2, ERBB3, ERBB4, PIK3CA,* and *PTEN*; and transcription of *AKT1, AKT2, AKT3, PTEN, PIK3CA, EGFR, ERBB2, ERBB3, ERBB4, NRG1, NRG2, HBEGF, BTC,* and *EREG* [26]. The TCGA-SKCM dataset is publicly available through the NIH/NCI Genomic Data Commons (GDC) portal [74]. The R statistical computing and graphics environment software [75] was used to reorganize the dataset. Statistical analyses were performed using GraphPad Prism [76] and Microsoft Excel [77].

### B. Cell lines and cell culture

The mouse C127 fibroblast cell line [78], the ψ2 ectropic recombinant retrovirus packaging cell line [79], and the PA317 amphotropic recombinant retrovirus packaging cell line [80] are generous gifts of Daniel DiMaio (Yale University). These cells were cultured essentially as described previously [63, 78–82]. The MEL-JUSO [50] and IPC-298 [49] human *BRAF* WT melanoma cell lines were obtained from DSMZ [83] (Braunschweig, Germany) and were cultured as recommended by the vendor. The MeWo [51] and SK-MEL-2 human *BRAF* WT melanoma cell lines were obtained from the American Type Culture Collection (ATCC - Manassas, VA) [84] and were cultured as recommended by the vendor. Cell culture media, sera, and supplements were obtained from Cytiva [85] (Marlborough, VA). G418 was obtained from Corning [86] (Corning, NY). Gene mutation and transcription data for the cell lines were obtained from the Broad Institute Cancer Cell Line Encyclopedia (CCLE) [46].

### C. Recombinant retroviruses

The recombinant amphotropic retroviruses LXSN [87], LXSN-ERBB4-WT [71], LXSN-ERBB4-K751M (LXSN-ERBB4-DN) [59, 65], LXSN-EGFR-WT [59, 71], LXSN-EGFR-K721A (LXSN-EGFR-DN) [59], LXSN-ERBB2-WT [59, 71], and LXSN-ERBB2-K753A (LXSN-ERBB2-DN) [59] were packaged using the ψ2 ecotropic retrovirus packaging cell line and the PA317 amphotropic retrovirus packaging cell line essentially as previously described [63, 82]. Briefly, because we wished to infect both murine and human cells, high titer recombinant amphotropic retrovirus stocks were required. Thus, the aforementioned recombinant retroviral plasmids were transfected using calcium phosphate into the ψ2 ecotropic retrovirus packaging cell line. Because the pLXSN recombinant retrovirus vector contains the neomycin resistance gene, stably transfected ψ2 cells were selected using G418. For each recombinant retrovirus construct, the colonies of stably transfected ψ2 cells were pooled and expanded. The conditioned medium of each resulting ψ2 cell line typically contains a low concentration (∼10^4 infectious units/mL) of recombinant ecotropic retrovirus. (Presumably this low titer is due to relatively inefficient packaging of retroviral genomes arising from transfected concatemerized plasmid DNA.) The PA317 amphotropic retrovirus packaging cell line was infected with each of these low-titer ecotropic retrovirus stocks. Stably infected PA317 cells were selected using G418 and these colonies were pooled and expanded. The conditioned medium of each resulting PA317 cell line typically contains a high concentration (∼10^6 infectious units/mL) amphotropic retrovirus. (Presumably this high titer is due to relatively efficient packaging of retroviral genomes arising from retrovirus infection.) These recombinant retroviruses were titered by infecting mouse C127 fibroblasts and selecting for stably infected cells using G418. Colonies of stably infected cells were manually counted and the number of G418-resistant colonies was divided by the volume of retrovirus used in the infection to determine the titer of each retrovirus stock.

### D. Clonogenic proliferation assays

C127, IPC-298, MEL-JUSO, MeWo, and SK-MEL-2 cells were infected with 500, 20000, 3000, 3000, and 20000 (respectively) infectious units (titered using C127 cells) of amphotropic retroviruses essentially as published [63–68]. Infected cells were selected using 800 ug/mL G418. The resulting colonies of G418-resistant cells were stained using Giemsa when distinct (typically 8-17 days later). Colonies were counted manually and plates were digitized for archival purposes. C127 infections served as controls for viral titer and clonogenic proliferation efficiency was calculated as previously described [63]. Briefly, we calculated the recombinant retroviral titer for each combination of cell line and virus. For each trial, we calculated the efficiency of clonogenic proliferation in the infected IPC-298, MEL-JUSO, MeWo, and SK-MEL-2 cells by dividing the recombinant retroviral titer in each of these cell lines by the corresponding recombinant retroviral titer in the C127 cell line. For each of the subject cell lines, the efficiency of the subject retroviruses was normalized to the efficiency of the LXSN vector control retrovirus in that same cell line. We report the average efficiency of clonogenic proliferation over a minimum of four independent trials. We used ANOVA to determine whether the efficiency of clonogenic proliferation of the *BRAF* WT melanoma cell lines infected with the subject retroviruses is significantly different from the efficiency of clonogenic proliferation of these cells infected with the vector control LXSN virus. We used a p-value threshold of <0.05 (1-tailed).

### E. Immunoprecipitation and immunoblotting

We performed ERBB4 immunoprecipitation and immunoblotting essentially as described [65, 67, 68, 71]. PA317 cell lines that had been stably infected with the LXSN vector control recombinant retrovirus, the LXSN-ERBB4-WT recombinant retrovirus, or the LXSN-ERBB4-DN recombinant retrovirus were grown to confluence. Approximately 2×10^7 cells were lysed on ice in 2 mL of ice-cold EBC lysis buffer (50 mM Tris, pH 7.4; 120 mM NaCl; 0.5% Igepal CA-630) supplemented with the protease inhibitor aprotinin (final concentration of ∼100 KIU/mL) and the protein phosphatase inhibitor sodium orthovanadate (final concentration of 1 mM). Centrifugation (14000 g*min at 4° C) cleared the lysates of intact nuclei and other debris. A Bradford assay was used to determine the protein concentration of the cleared lysates.

ERBB4 was immunoprecipitated from 1 mg of lysate diluted to a final concentration of 1 mg/mL. We used 10 uL of the anti-ERBB4 mouse monoclonal antibody SC-8050 (Santa Cruz Biotechnology) and 30 uL of a 1:1 suspension (in lysis buffer) of Protein A Sepharose Beads (17-0780-01; Sigma-Aldrich). The immunoprecipitation reactions were incubated at 4° C for 2 hours on a rocking platform. Afterward, the beads were collected by centrifugation (14000 g*min at 4° C) and washed three times with 500 uL ice-cold NET-N (20 mM Tris, pH 8.0; 100 mM NaCl; 1 mM EDTA, pH 8.0; 0.5% Igepal CA-630) supplemented with the protease inhibitor aprotinin (final concentration of ∼100 KIU/mL) and the protein phosphatase inhibitor sodium orthovanadate (final concentration of 1 mM).

Following the last wash step, the precipitated proteins were eluted from the beads by adding 80 uL of protein sample buffer (4% SDS; 0.1 M DTT; 0.3 M Tris, pH 7.4; 20% glycerol; 0.04% bromphenol blue; 10% 2-mercaptoethanol). The samples were mixed using a vortex mixer and boiled for 5 minutes. The beads were collected by centrifugation (14000 g*min at room temperature) to permit retrieval of the eluted proteins.

The eluted proteins were resolved by SDS-PAGE using a 7.5% acrylamide gel and electrotransferred (200 mA for >12 hours) to 0.2 um PVDF in a wet tank transfer system (TE-42; Hoefer) containing a transfer buffer consisting of 20% methanol, 25 mM Tris base, 192 mM glycine, 0.09375 % SDS, and 0.01 % sodium orthovanadate. The blot was rinsed three times for 5 minutes at room temperature using TBS-T (10 mM Tris, pH 7.4; 9 g/L NaCl; 0.05% Tween-20), then blocked for two hours at room temperature on a rocking platform using 5% non-fat dried milk in TBS-T. If the blot was to be probed using an anti-phosphotyrosine antibody, the blot was blocked for two hours at room temperature on a rocking platform using 5% BSA and 0.01% sodium azide in TBS-T.

The ERBB4 blots were incubated for two hours at room temperature on a rocking platform with an anti-ERBB4 rabbit monoclonal antibody (4795S – Cell Signaling Technology) diluted 1:1000 in 5% non-fat dried milk in TBS-T. The blots were washed 5 times with TBS-T for 5 minutes at room temperature on a rotating platform. The blots were then probed with a goat anti-rabbit antibody conjugated with horseradish peroxidase (31460 - ThermoFisher). Antibody binding was detected using enhanced chemiluminescence (Cytiva) and documented using a BioRad Chemidoc MP imaging system.

The phosphotyrosine blots were incubated for two hours at room temperature on a rocking platform with the anti-phosphotyrosine mouse monoclonal antibody 4G10 (made in-house from hybridoma PTA-6854; ATCC) diluted empirically in 5% BSA and 0.01% sodium azide in TBS-T. The blots were washed 5 times with TBS-T for 5 minutes at room temperature on a rotating platform. The blots were then probed with a goat anti-mouse antibody conjugated with horseradish peroxidase (31430 - ThermoFisher). Antibody binding was detected using enhanced chemiluminescence (Cytiva) and documented using a BioRad Chemidoc MP imaging system.

### F. Anchorage-independent proliferation assay

We performed this assay essentially as described [66, 78, 88]. Briefly, for each MEL-JUSO stably infected cell line, 20,000 cells were seeded in a 60 mm cell culture dish in a semi-solid medium consisting of RPMI-1640 supplemented with 10% fetal bovine serum, 500 ug/mL G418, and 0.3% low-melting point (LMP) agarose. Additional LMP agarose medium was added to the plates every four days to prevent the cultures from drying out. Fourteen days after seeding, nine randomly selected microscopic fields were digitally photographed. We used NIH Image J [89] to measure the diameter of each anchorage-independent colony of cells. Approximately 8-12 colonies were found in each photograph. Thus, for each trial, approximately 100 colonies were measured for each cell line. We pooled the data from three independent trials, resulting in approximately 300 colonies for each cell line. A t-test was performed to determine whether differences in average colony size are statistically different.

## Discussion

### A. Elevated ERBB4 signaling appears to drive BRAF WT melanomas

Our data suggest that approximately 12% of *BRAF* WT melanomas exhibit elevated endogenous *ERBB4* transcription, which presumably results in elevated ERBB4 signaling. Elevated endogenous ERBB4 expression is significantly correlated with *NF1*/*RAS* gene tumor driver mutations. Consequently, we postulated that elevated ERBB4 signaling cooperates with elevated RAS signaling to drive *BRAF* WT melanomas. We tested this hypothesis by measuring the effects of exogenous expression of the WT *ERBB4* allele or an *ERBB4* DN mutant allele on the clonogenic proliferation of BRAF WT melanoma cell lines.

WT *ERBB4* stimulates clonogenic proliferation of IPC-298, MEL-JUSO, MeWo, and SK-MEL-2 *BRAF* WT melanoma cells. Moreover, the *ERBB4* DN (K751M) mutant allele inhibits clonogenic proliferation of these same cell lines. These results indicate that ERBB4 is sufficient and necessary for the clonogenic proliferation of these *BRAF* WT melanoma cell lines.

Numerous *ERBB4* mutant alleles have been found in melanomas and other human tumor samples [7]. The data presented here suggest that some of these mutants exhibit a gain-of-function phenotype that enables them to serve as tumor drivers. We will test that hypothesis.

### B. RAS pathway mutations and elevated heterotypic ERBB4 signaling suggest strategies for treating BRAF WT melanomas

It is commonly accepted that ERBB4 homodimers inhibit cell proliferation, whereas ERBB4-EGFR or ERBB4-ERBB2 heterodimers stimulate cell proliferation [7]. Likewise, published data [53, 59, 71] and data shown here suggest that elevated heterotypic ERBB4 signaling (by ERBB4-ERBB2 or ERBB4-EGFR heterodimers) causes increased PI3K signaling, which cooperates with elevated RAS signaling to drive the proliferation of *BRAF* WT melanomas (**Figure 6**). Thus, we predict that ERBB4-dependent, *BRAF* WT melanomas will respond to a combination of a PI3K inhibitor with a MEK inhibitor. However, given the toxicity of PI3K inhibitors [90, 91], combining a MEK inhibitor with an anti-EGFR or anti-ERBB2 agent may be a more effective treatment of ERBB4-dependent, *BRAF* WT melanomas than the combination of a MEK inhibitor with a PI3K inhibitor. We will test these predictions.

**Figure 6.**
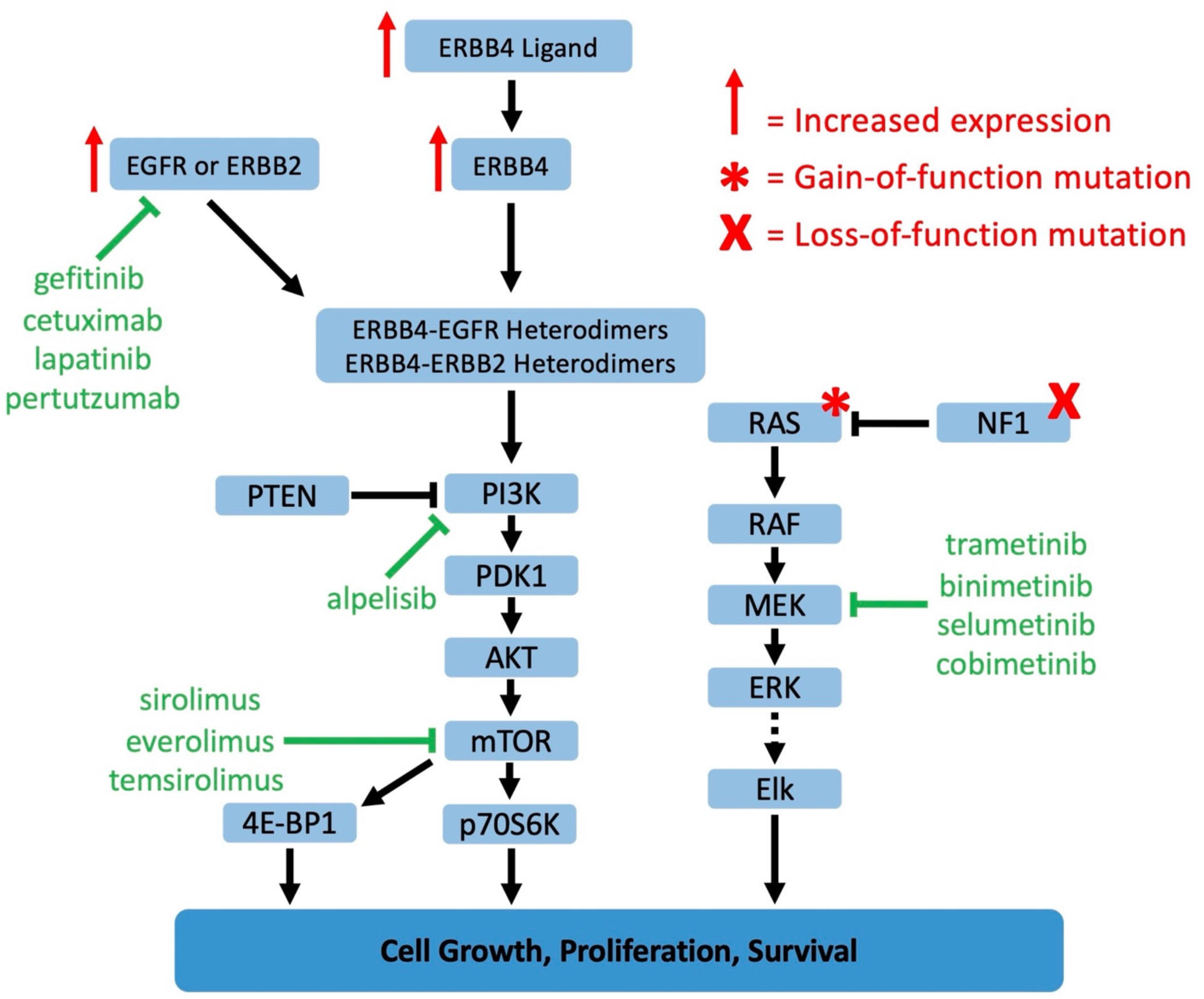
We hypothesize that signaling by ERBB4-EGFR or ERBB4-ERBB2 heterodimers can be stimulated by ERBB4 ligands, ERBB4 overexpression, or ERBB4 mutations, resulting in increased signaling by the PI3K pathway, which cooperates with elevated RAS/MAPK pathway signaling to drive the proliferation of *BRAF* WT melanomas.

### C. Potential mechanisms of specificity in *ERBB4*-dependent *BRAF*-WT melanomas

Here we demonstrate that *ERBB2*, but not *EGFR*, is necessary for clonogenic proliferation of MEL-JUSO cells. This specificity may not be shared by the IPC-298, MeWo, and SK-MEL-2 cell lines, as *EGFR* but not *ERBB2* may be necessary for the clonogenic proliferation of these other *BRAF* WT melanoma cell lines. Furthermore, differences in the role that *EGFR* and *ERBB2* play in the four *ERBB4*-dependent, *BRAF* WT melanoma cell lines may account for the gender disparity observed in *BRAF* WT melanoma outcomes (**Table 2c**), particularly since the ERBB4 cytoplasmic domain can translocate to the nucleus and possesses a motif that enables interactions with steroid hormone receptor co-activators [7]. Finally, it would not be surprising if some *ERBB4* gain-of-function mutant alleles found in *BRAF* WT melanoma samples and elsewhere [7] function through potentiating ERBB4 heterodimerization with ERBB2 but not EGFR or vice versa. Clearly, much experimentation lies ahead to address these pressing questions and to determine whether the observations made using *ERBB4*-dependent, *BRAF* WT melanoma cell lines can be translated into advances in the treatment of *BRAF* WT melanomas.

## Data Availability

All data produced in the present study are available upon reasonable request to the authors.

https://portal.gdc.cancer.gov/projects/TCGA-SKCM

